# Patient’s age impact on stroke risk and intracranial aneurysm occlusion - SCENT Flow Diverter trial on large and giant aneurysms

**DOI:** 10.1101/2024.12.12.24318961

**Authors:** Ajay K. Wakhloo, Paul Jenkins, Philip M. Meyers, Alexander L. Coon, Peter T. Kan, Ajit Puri, Joost de Vries, Giuseppe Lanzino, Mark Bain, Koji C. Ebersole, Babu G. Welch, Aclan Dogan, Pascal Jabbour, J Mocco, Adnan H. Siddiqui, Quill Turk, Philipp Taussky, Ricardo A. Hanel, the SCENT Investigators

**Affiliations:** Department of Radiology, Tufts University School of Medicine, Boston, MA; Statistical Consultant; Departments of Radiology and Neurological Surgery, Columbia University, New York, NY; Carondelet Neurosurgical Institute of St. Joseph’s Hospital, Tucson, AZ; Department of Neurosurgery, University of Texas Medical Center, Galveston, TX; Division of Neurointerventional Radiology, UMASS Chen School of Medicine, Worcester, MA; Department of Neurosurgery, Radboud, University of Nijmegen, Nijmegen, The Netherlands; Department of Neurosurgery, Mayo Clinic, Rochester, MN; Department of Neurosurgery, Cerebrovascular Center Cleveland Clinic, Cleveland, OH; Department of Neurosurgery, University of Kansas Health Systems, Kansas City, KS; Department of Neurosurgery, University of Texas Southwestern Medical Center, Dallas, TX; Department of Neurosurgery, Oregon Health & Science University, Portland, OR; Department of Neurosurgery, Thomas Jefferson University Hospital, Philadelphia, PA; Department of Neurosurgery, Mt. Sinai Medical Center, New York, NY; Department of Neurosurgery, SUNY University at Buffalo, Buffalo, NY; Department of Radiology, Medical University of South Carolina, Charleston, SC; Department of Neurosurgery, Beth Israel Deaconess Hospital, Harvard Medical School, Boston, MA; Lyerly Neurosurgery, Jacksonville, FL

## Abstract

**Importance:** The multicenter, prospective, single-arm, non-randomized SCENT trial on flow diverter (FD) treatment for intracranial aneurysms (IA) was analyzed for patients’ age and IA characteristics impacting stroke and occlusion rates over 5 years.

**Design:** The impact on major ipsilateral stroke and IA occlusion was studied by stratifying age as ≤ 65 years versus >65 years. Product-limit (Kaplan-Meier) estimates of time to both endpoints, stratified by age group, were created. Univariate predictors of time to stroke were identified by including candidate variables in univariable proportional hazards regression models. Those variables found to be significant (p<0.10) at the univariate level were entered into a multivariable survival model to identify independent predictors. The stepwise selection produced a final reduced model with a significance level to both enter and stay set at 0.05.

**Findings:** Of 180 patients with 180 large or giant IA enrolled in the modified intention-to-treat cohort, 119 subjects were ≤ 65, while 61 patients were > 65 years old. When parent artery stenosis and IA size were entered into a multiple-stepwise survival model, only stenosis remained as an independently significant predictor of time to stroke. At 3-year follow-up, there were a total of 23 strokes (12.8%), with 11 occurring in subjects ≤ 65 years; there was a greater risk for seniors (HR1.96, 95% CI 0.83-4.78). Four patients (4/180; 2.2%) experienced aneurysm rupture within the first week post-treatment, with 3 being ≤ 65 and the fourth aged 66 years. No new strokes were reported between 3 and 5 years. Complete IA occlusion rates for seniors were 60.0% (33/55), 67.6% (25/37), and 85.7% (18/21) at 12, 36, and 60 months, respectively, as compared to 79.1% (87/110), 82.5% (66/80), and 91.8% (56/61) for younger subjects. The time to complete IA occlusion was shorter in younger patients (HR1.53, 95% CI 1.07-2.19). Five subjects (2.8%) underwent retreatment, 2 in 60-year-old patients, and one each aged 64, 70, and 75.

**Conclusions and Relevance:** Age > 65 and parent artery stenosis are related to an increased risk of major ipsilateral stroke in patients with intracranial aneurysms treated with a flow diverter. Age > 65 is also predictive of increased time to and incomplete healing. With demographic shifts, future treatments need to focus on expedited and improved healing.

**Trial Registration:** https://www.clinicaltrials.gov NCT01716117

**KEY POINTS:** *Question:* What key variables, including patients’ age and intracranial aneurysm (IA) characteristics, determine procedural stroke risks in subjects treated for large or giant IA with flow-diverting stents?

*Findings:* Aneurysm size and parent artery stenosis impacted occlusion rates and stroke risk, respectively. Over a 5-year observation period, the senior population had a significantly lower occlusion rate and higher risk for stroke, while the younger population was at higher risk for early aneurysm bleed following treatment.

*Meaning:* With demographic shifts and a higher senior population being treated for IA, the stroke risk and incomplete occlusion rates need to be discussed with the patient before treatment.

## INTRODUCTION

Aging has a significant negative impact on peri-and postoperative risk and delaying postoperative wound healing.^1^ Analogously, in cardio-, peripheral-, and cerebrovascular settings, higher age-related morbidity and mortality, as well as delayed and incomplete healing of intravascular implants, such as stents and coils, have been reported.^2–4^ For over a decade, flow diverters (FDs) have been used to manage intracranial aneurysms (IA) and have continuously grown to mainstream endovascular treatment for a subset of IA with high rates of progressive and complete obliteration.^5,6^ Due to a demographic shift and widespread use of non-invasive imaging modality, a larger senior population harboring IA is being treated.^7^ We studied the patient’s age and aneurysm characteristics that may affect the peri- and postprocedural and long-term stroke risk, as well as a complete and definite occlusion of IA treated with FD. A substudy of the SCENT (Surpass Intracranial Aneurysm Embolization System Pivotal Trial to Treat Large or Giant Wide Neck Aneurysms) was performed, the largest controlled trial on FD for IA to date.^8,9^ Possible mechanisms for our observations will be discussed.

## METHODS

SCENT was a multicenter, prospective, single-arm, and, for ethical reasons, a non-randomized trial with an independent clinical event committee and a core lab adjudicating the safety and efficacy of the Surpass Streamline FD in clinical practice. Control groups were historic. Preoperative assessment was performed according to the trial protocol (https://www.clinicaltrials.gov NCT01716117). Detailed study design, enrollment, and data have been published previously.^8,9^ Study enrollers were from 25 centers in the US and one center in Europe. Institutional review board approval of the investigational device exemption study protocol was obtained at each participating center. Written informed consent was obtained from each patient before participation in SCENT.

### Endovascular procedure

Procedural preparation included local standard of care preoperative evaluation and achievement of high-level antithrombotic, dual antiplatelet therapy. The dual antiplatelet therapy protocol in SCENT required the administration of aspirin 75 to 325 mg orally per day and clopidogrel 75 mg orally per day for at least 5 days before the investigational procedure. Following device implantation, aspirin 75 to 325 mg orally daily was recommended to be continued for the rest of the patient’s life. In contrast, clopidogrel 75 mg orally per day or its pharmacological equivalent was required for at least 6 months. The antiplatelet activity was assessed according to the standard of care at the enrolling site using genetic testing or platelet aggregometry. Platelet inhibition between 30% and 90% was considered therapeutic for the procedure. Alternatively, genetic testing using CYP2C19 allele testing was used by some sites to predict individual patient responsiveness to clopidogrel instead of platelet aggregometry. ^10^ A trained site investigator performed each procedure. The trial protocol allowed placement of 1 to 2 devices, preferably a single device, unless a second was needed to improve proximal wall apposition or if the first device did not completely cover the aneurysm neck. Adjunct coil embolization of the aneurysm was not allowed in the SCENT study.

### Study device, procedure, protocol, and follow-up

Subjects underwent independent neurological evaluation 12 to 36 hours post-procedure and at 1 month, 6 and 12 months, 3, and 5 years. The composite primary effectiveness endpoint included three components per FDA requirements: complete aneurysm occlusion (Raymond-Roy (RR) class 1 parent artery stenosis, and no retreatment within 12 months.^11^ The protocol specified that the primary safety endpoint was the rate of major ipsilateral stroke (increase in National Institutes of Health Stroke Scale [NIHSS] score of ≥4) or neurological mediated death within 12 months. Additional post hoc analysis of the primary safety endpoint was performed at the request of the FDA using a modified endpoint definition of disabling stroke or neurological death, wherein disabling stroke was defined as a modified Rankin Scale (mRS) score of ≥3 at a minimum of 90 days post-stroke event. [Stryker Neurovascular. Surpass Streamline™ Flow Diverter Directions for Use. Food and Drug Administration. NV00024766 rev 16. https://www.accessdata.fda.gov/cdrh_docs/pdf17/P170024C.pdf. Accessed September 28, 2018] A subject lost to follow-up was defined as a subject who could not be located after multiple attempts and who missed their follow-up appointments or was unable to follow up due to death.

### Statistical analysis

The two primary endpoints for analyses were the time from procedure to 1) major ipsilateral stroke and 2) complete aneurysm occlusion based on Raymond-Roy classification. While the first of these two endpoints could occur at any time, the second, Raymond-Roy class, was assessed at post-procedure, 6 and 12 months, 3 and 5 years. The time to major ipsilateral stroke was right-censored at 1-year post-procedure, and the time to Raymond-Roy Class 1 was right-censored at 5 years. The effect of age on time on both major ipsilateral stroke and Raymond Class 1 was examined by stratifying age as ≤65 years versus >65 years. Product-limit (Kaplan-Meier) estimates of time to both endpoints, stratified by age group, were then created. Univariate predictors of time to major ipsilateral stroke were identified by including candidate variables in univariable proportional hazards regression models. Those variables found to be significant (p < 0.10) at the univariate level were entered into a multivariable survival model to identify independent predictors of this outcome. Stepwise selection produced a final reduced model with a significance level to enter and stay at 0.05. All analyses were performed using SAS 9.4 (SAS Institute Inc, Cary, NC).

## RESULTS

A total of 180 patients with 180 aneurysms were enrolled in the modified intention-to-treat cohort. Almost twice as many patients treated were 65 years or younger (n=119), and 61 patients were > 65 years. Based on the imaging core lab assessment, 15 aneurysms (8.3%) were fusiform, 164 (91.1%) saccular IA, and 1 (0.6%) blister aneurysm. The mean aneurysm size was 14.2 mm (SD 6.1). Thirteen (7.4%) aneurysms were giant (≥25 mm). Fifty-eight (32.2%) aneurysms were in the supraclinoid and distal (including posterior communicating artery) segments of the internal carotid artery. The mean procedure duration was 53.6 minutes. The device was successfully implanted in 97.8% of patients, with a mean of 1.1 devices per patient.

### Complications

The twelve-month major ipsilateral stroke or neurological death rate was 11.1% [(20/180); 95% CI, 6.9–16.6]. Results of the univariates analyses of time to major ipsilateral stroke are shown in table 1 and figure 1. Significant univariate predictors of this outcome were pre-existing parent artery stenosis (HR 1.03, 95% CI 1.00 – 1.06, p=.031), aneurysm dome depth (HR 1.07, 95% CI 1.00 – 1.15, p=.049), and dome height (HR 1.06, 95% CI 1.00 – 1.12, p=.038). A near-significant effect was also seen for aneurysm size (HR 1.06, 95% CI 1.00 – 1.12, p=.071). These results are shown in a Forrest Plot (Figure 1). At the 3-year follow-up point, there were a total of 23 major ipsilateral strokes (12.8%), with 11 (47.8%) occurring in subjects aged 65 and younger. No new cases of major ipsilateral stroke occurred between the 3-year and 5-year time points. Four patients (4/180; 2.2%) experienced aneurysm ruptured within the first week post-treatment, with 3 of these being ≤65 and the fourth aged 66 years. No further aneurysm ruptures occurred during the 5-year follow-up. New serious adverse events at 3 and 5 years were noted in 15.4% (23/149) and 9.8% (12/123) of patients, respectively. Five neurological deaths were seen within one year, and one additional neurological death occurred beyond the 5-year follow-up window at 1,849 days. Of the five neurological deaths occurring within the 5-year window, two occurred in subjects aged more than 65 years.

**Figure 1:**
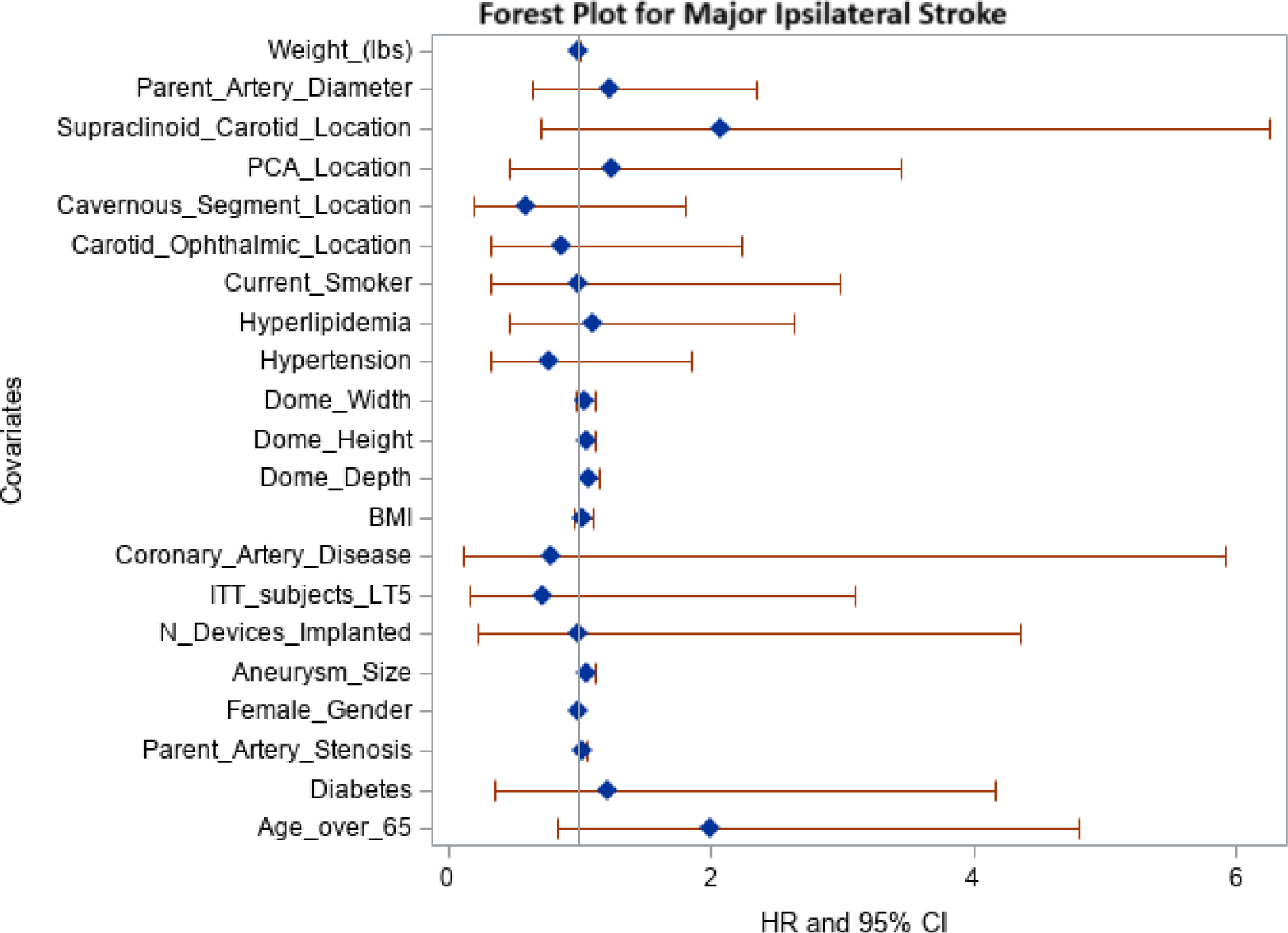
Forest plot of univariate predictors of time to major ipsilateral stroke.

**Table 1.** Univariate Predictors of Time to Major Ipsilateral Stroke.

| Covariate | Hazard Ratio (95% CI) | p-value |
| --- | --- | --- |
| Age Greater than 65 | 1.96 (0.83 – 4.78) | .122 |
| Diabetes | 1.22 (0.36 – 4.17) | .748 |
| Parent Artery Stenosis | 1.03 (1.00 – 1.06) | .031 |
| Female Gender | 1.00 (1.00 – 1.00) | .575 |
| Aneurysm Size | 1.06 (1.00 – 1.12) | .071 |
| Number of Devices Implanted | 1.00 (0.23 – 4.36) | .996 |
| Site has <5 ITT subjects | 0.72 (0.17 – 3.10) | .660 |
| Coronary Artery Disease | 0.79 (0.11 – 5.91) | .820 |
| Body Mass Index | 1.02 (0.96 – 1.10) | .504 |
| Dome Depth | 1.07 (1.00 – 1.15) | .049 |
| Dome Height | 1.06 (1.00 – 1.12) | .038 |
| Dome Width | 1.04 (0.98 – 1.12) | .221 |
| Hypertension | 0.77 (0.32 – 1.85) | .558 |
| Hyperlipidemia | 1.10 (0.46 – 2.63) | .839 |
| Current Smoker | 1.00 (0.33 – 2.99) | .999 |
| Carotid Ophthalmic Location | 0.86 (0.33 – 2.23) | .752 |
| Cavernous Segment Location | 0.60 (0.20 – 1.80) | .364 |
| PCA Location | 1.25 (0.46 – 3.45) | .660 |
| Supraclinoid Carotid Location | 2.08 (0.70 – 6.25) | .189 |
| Parent Artery Reference Diameter | 1.23 (0.65 – 2.34) | .528 |
| Weight (lbs) | 1.00 (0.99 – 1.01) | .575 |

When parent artery stenosis, dome depth, dome height, and aneurysm size were entered into a multiple-stepwise survival model, only parent artery stenosis remained as an independently significant predictor of time to major ipsilateral stroke. Figure 2 shows Kaplan-Meier curves for time to major ipsilateral stroke stratified by age group. There was a trend of greater risk for the older age group, but it did not reach significance (HR 1.96, 95% CI 0.83 - 4.78, p=.122). Similarly, supraclinoid carotid artery aneurysm location showed a trend toward higher risk for ipsilateral stroke (HR of 2.08, 95% CI 0.70-6.25, p=.189). A balloon angioplasty following FD placement to improve wall apposition was used in 123 patients; one subject was treated with an additional stent. Among the 61 subjects > 65, 38 (62.3%) had post-deployment angioplasty, and in the ≤ 65 population, angioplasty was carried out in 67 subjects (56.3%, p=.440).

**Figure 2:**
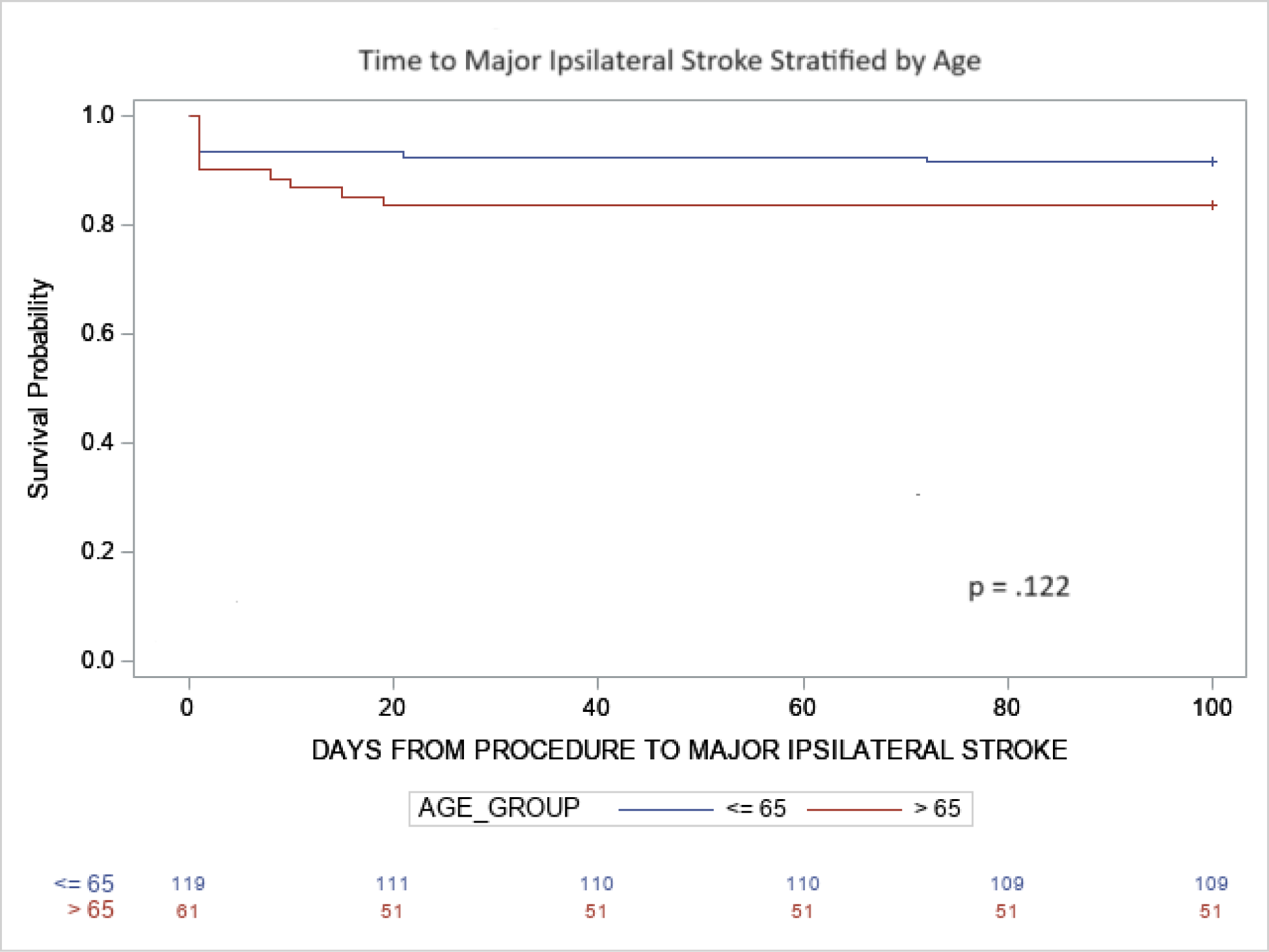
Kaplan-Meier curve for time to major ipsilateral stroke stratified by age.

### Aneurysm occlusion rates

The primary effectiveness outcome was the percentage of subjects with 100% aneurysm occlusion (RR class I) without significant stenosis of the treated segment of the internal carotid artery. The twelve-month RR I rate was 72.7% [(120/165); 95% CI, 65.9–79.5%]. In 15 patients (8.3%), angiograms were unavailable at 12 months to assess the occlusion rates. The three-year and 5-year occlusion rates increased to 77.8% (91/117) and 90.2% (74/82) respectively. In the senior population, RR I was achieved in 50% (27/54), 60.0% (33/55), 67.6% (25/37), and 85.7% (18/21) at 6,12, 36, and 60 months respectively (Figure 3). At the same time points, in subjects ≤65 years, the RR1 rates were 73.0% (81/111), 79.1% (87/110), 82.5% (66/80), and 91.8% (56/61). The time to complete aneurysm occlusion was shorter in the younger age stratum (HR 1.53, 95% CI 1.07 – 2.19, p=.020). Five aneurysms (2.8%) were retreated in the study. These retreatments occurred in 2 subjects aged 60, and one each aged 64, 70, and 75.

**Figure 3:**
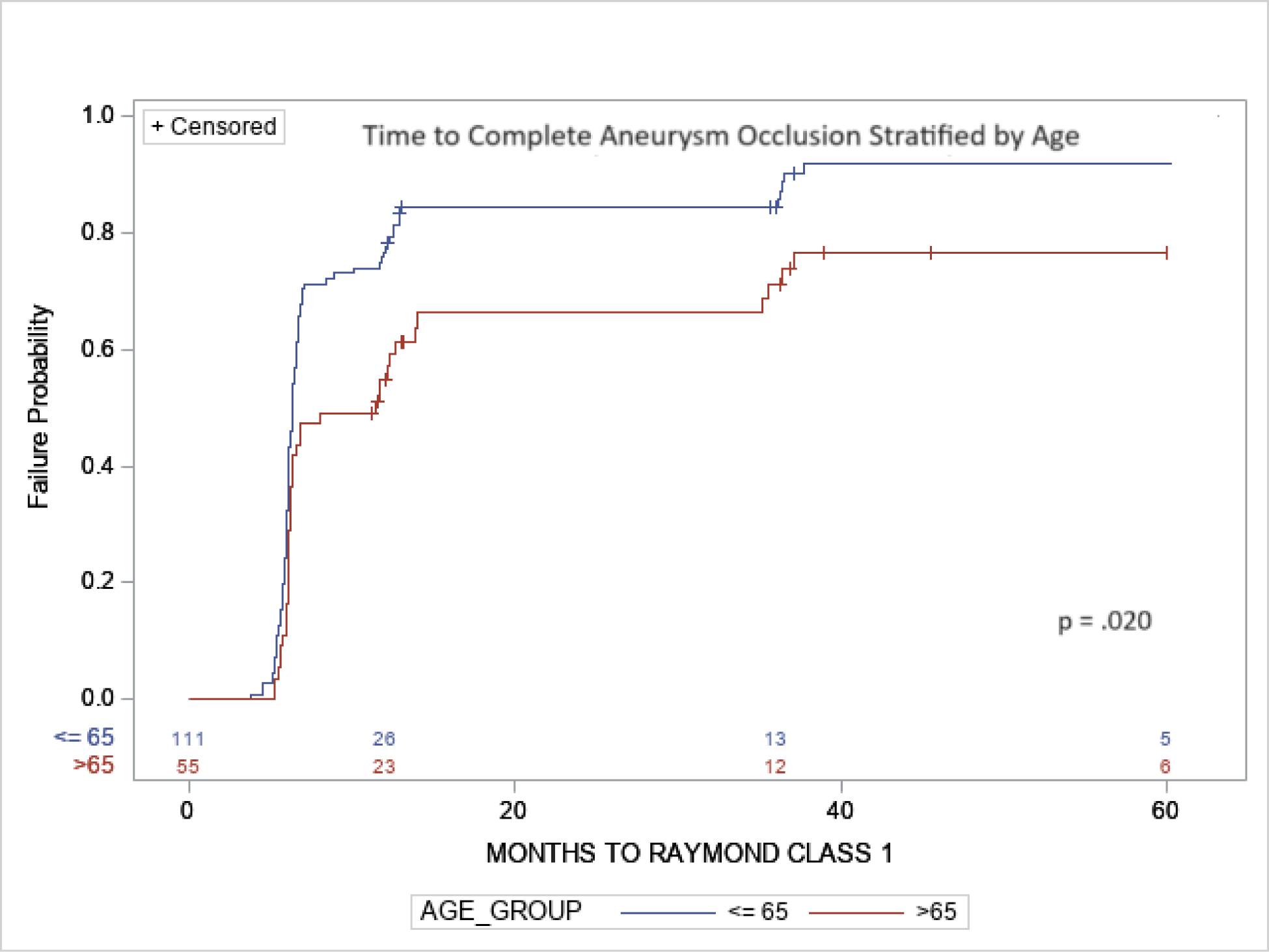
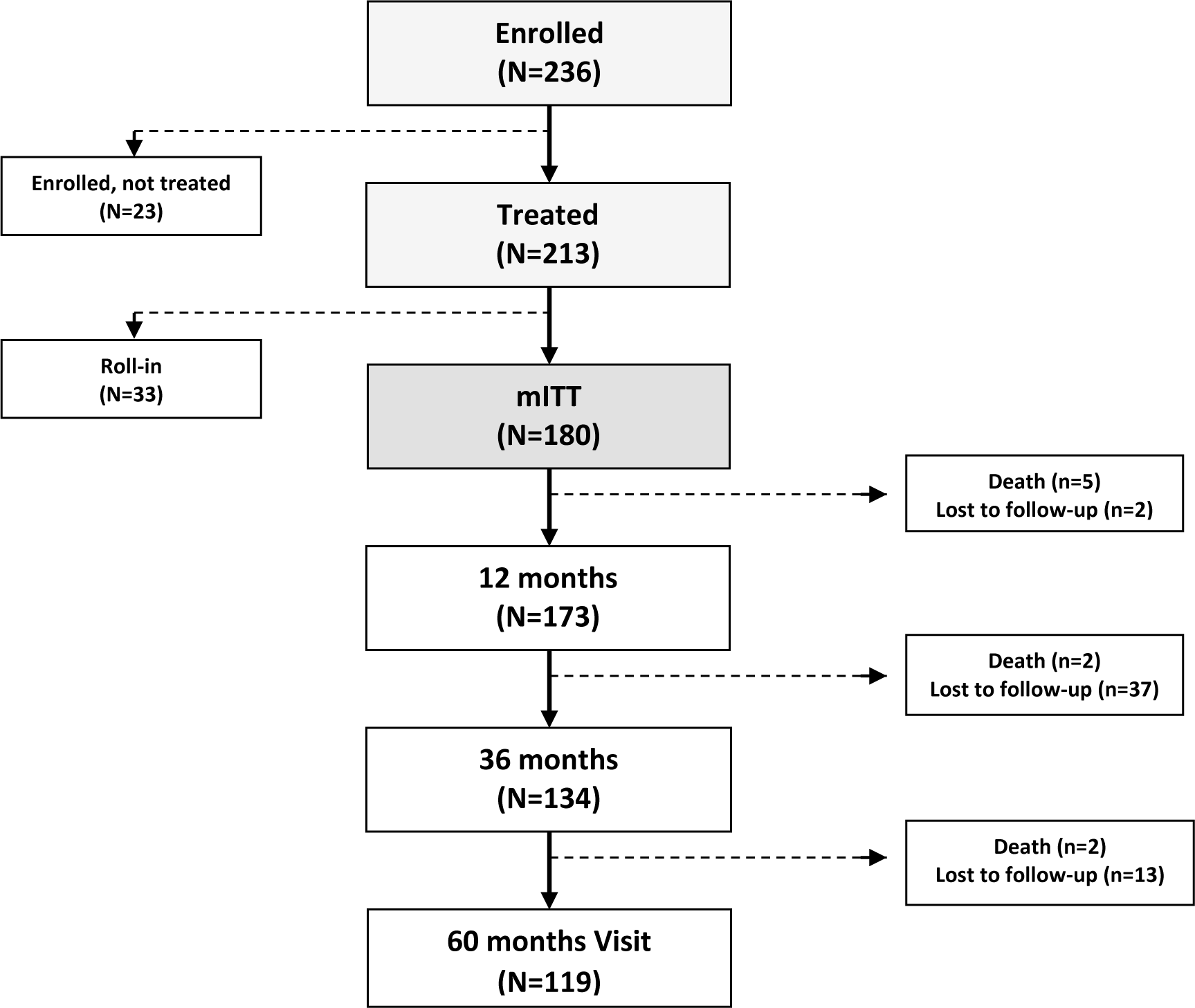
Kaplan-Meier curves for time to complete aneurysm occlusion (Raymond-Roy Class 1) stratified by age group.

The most common medical comorbidities frequently associated with age included hypertension, hyperlipidemia, headache, depression, and anxiety. Twenty-six (14.4%) patients had a history of prior aneurysmal subarachnoid hemorrhage (SAH). Forty-one procedures (i.e., intracranial clipping, coiling, and stent placement) to treat another (non-target) aneurysm had previously occurred in 27 (15.0%) patients. Thirty (16.7%) patients had undergone prior coiling (26, 14.4%) or surgical clipping (4, 2.2%) of target aneurysms. There was a high percentage of current (20%) or prior (44%) smokers. Sixteen (8.9%) had a history of prior stroke.

## DISCUSSION

Although the overall annual risk for aneurysm rupture is low at 0.7% - 0.95%, large and giant IA have a poor natural history after diagnosis. A recent study from Japan showed a hazard ratio for 10 to 24 mm size of 9.09 (95% CI, 5.25 to 15.74) and 25 mm or larger aneurysms, 76.26 (95% CI, 32.76 to 177.54) requiring a treatment. ^12,13^ The study evaluated 6697 unruptured IA and found aneurysms 7 mm or larger in 39.7% of patients ≥80 years compared to 21.4% of patients 50–59 years. The risk of IA rupture increases with age, ethnicity, aneurysm size, location, history of previous subarachnoid hemorrhage from another aneurysm, and arterial hypertension, which are significant risk factors.^14^

As shown in various prospective controlled studies, flow diverters represent a paradigm shift in managing large and giant IA with acceptable periprocedural morbidity and mortality and high long-term success.^5,6,8,9^ While the proportion of individuals ≥65 years or older in the general population is predicted to more than double from 8% in 2010 to 21% in 2050, the health management of this subpopulation remains controversial.^15^ Advanced age is a risk factor for peri-procedural morbidity and mortality after coiling and clipping of IA. However, limited information is available for patients treated with flow diverters. For subjects > 60 years, Pierot et al. found in the AETNA study that coiling for IA was associated with morbidity and mortality of 3.8% and 2.3%, respectively. In comparison, the risk decreased to 1.2% for mortality and 1.2% for morbidity in the population £ 60 years.^2^ Long-term data on AETNA have not been published. Yet, with continuous technology improvements as well as global demographic shifts and changes in life expectancy, an increased number of senior patients are being treated endovascularly for ruptured and unruptured IA.^16,17^ Previous studies noted a significant increase in the proportion of patients undergoing endovascular coiling between 2001 and 2009 for all age groups.^7^ For both IA-clipped and coiled patients, mortality and the proportion of patients discharged to long-term facilities increased with age. Overall mortality for patients clipped and coiled decreased modestly for all age groups, and overall proportions of patients discharged home increased modestly (p=.01), except for patients older than 80.

### Ischemic events

Previous self-adjudicated reports from single and multicenter observational studies have found in the senior population an increased risk of thromboembolic ischemic stroke and impaired IA healing following FD treatment.^18^ We performed a subgroup analysis of currently the most extensive prospective controlled study on FDs for large and giant IA. Due to a small number of events, only aneurysm size and pre-existing parent artery stenosis remained as an independently significant predictor of time to major ipsilateral stroke. Our study also found a trend toward higher risk for an ipsilateral stroke for supraclinoid carotid artery aneurysms, most likely due to increased manipulation of the device delivery system. Increased age, although not significant, most likely due to the small sample size, was associated with increased risk for ischemic events. Several factors may contribute to these findings, i.e., hardening and calcifying of the aortic arch and cerebrovascular system and increased plasma concentration of procoagulant factors and platelet activity.^19^

Other risk factors include peri-aneurysmal vascular environment, arterial access, and increased tortuosity, especially of the internal carotid artery between the posterior genu of the siphon and supraclinoid segment, well defined on non-invasive imaging. They should be considered in pre-operative planning.^20^ Interestingly, in our study, we observed early rupture following FD treatment, most commonly in younger subjects. Early rupture is proposed to be related to an increased inflammatory response to a large amount of intra-aneurysmal clot forming following FD placement combined with dual antiplatelet management required for the implant.^21^ In experimental aneurysms, genes related to inflammation, i.e., tumor necrosis factor α and monocyte chemoattractant protein 1, were upregulated when treated with FD compared to coiled aneurysms, and enzymatic activity of active matrix metalloproteinase 9 was higher as well.^22^ An overshooting initial inflammatory response in younger subjects to the implant involving immune cells, such as macrophages, which release signaling molecules, may explain the breakdown of the vessel wall via smooth muscle cell expression of matrix metalloproteinases.^21^

### Occlusion rates

Pierrot et al. found, in the AETNA trial, complete aneurysm occlusion following coiling for subjects > 60 in 47.1%, while a complete occlusion in patients < 60 was seen in 65.2%.^3^ In another large single-center stent-assisted coiling study, investigators found, in univariate analysis, aneurysm size (>10 mm), increasing age > 65 years, and aneurysm location to be negative predictors of immediate occlusion. In univariate analysis, predictors of recanalization on follow-up were older age (> 65 years) and aneurysm size.^4^

Flow Diverters uncouple momentum and mass transfer from the parent artery into the aneurysm, creating flow reduction within the aneurysm, activation of the clotting cascade, and formation of an intra-aneurysmal clot formation with secondary scarring of the aneurysm pouch and endothelial coverage of FDs. This finally leads to progressive obliteration of the aneurysm from the parent artery.^23^ Our study shows that the time to complete IA occlusion is shorter in younger while older patients require more frequent early retreatment. On the other hand, the younger population has a higher risk for early spontaneous aneurysm rupture following FD treatment. Potential mechanisms for delayed aneurysm occlusion include a reduction in the age-related number of circulating and dormant precursor cells, the capability of dedifferentiation and proliferation, reduced motility of precursor cells, and genetic and epigenetic factors, which participate in endothelial reparative mechanisms.^24^ Nitric oxide (NO), which has vasodilatory and anti-inflammatory effects, promotes the migration and proliferation of endothelial cells and contributes to vascular remodeling.^25^

Furthermore, parent artery stenosis may affect a proper wall apposition of the FD, leading to delayed or incomplete endothelization and occlusion as previously described in preclinical models using high-frequency optical coherence tomography (OCT).^26^ Smaller cohort studies, previous studies did not find evidence that age is a risk factor for healing. Still, they found aneurysm size and gender, as well as incorporated branch vessels, to be strong predictors for incomplete healing and potentially requiring retreatment.^27^ A recent systematic review analyzing 24 mostly retrospective and a few single-center prospective studies out of 1184 screened publications found scarce evidence for predictors for IA occlusion following treatment with FDs.^28^ However, the literature suggests that the absence of branch involvement, younger age, and aneurysm size have the highest impact on aneurysm occlusion rates.

### Future work

Future work will need to focus on improving the wall apposition of FDs and enhancing and expediting endothelial repair and aneurysm occlusion rates by enhancing the proliferation and differentiation of progenitor cells that may be affected by aging. As a recent review paper summarized, newer implants that promote neointimal growth and decrease thrombogenicity via passive surface or active drug modifications are being studied.^29^ Preliminary preclinical studies in an experimental aneurysm model showed that Low Laser Light Therapy (LLLT, aka Photobiomodulation) delivered in situ in conjunction with FDs expedites complete healing of the aneurysm and endothelization of the implant.^30^ Photobiomodulation may be beneficial in upregulating reparative function by stimulating various types of stem cells to proliferate and differentiate.^31–33^ The work mechanisms are based on the electronic excitation of CuA and CuB chromophores in cytochrome c oxidase molecule. As a result, the cytochrome c oxidase molecule’s redox status and functional activity is modulated.^34^

Other investigators have proposed, in a rabbit aneurysm model, CD31-mimetic coated FD, to improve aneurysm occlusion rates and early endothelialization of the implant.^35^ In a mouse model of saccular aneurysm, monocyte chemoattractant protein 1 (MCP-1), osteopontin, and interleukin 10 (IL-10) have shown an increased aneurysm healing when locally delivered on the surface of coils.^36–39^ Recently, Laurent et al. published their results using dual-coated coils to target the inflammatory pathway of aneurysm healing.^40^ These efforts may not only eliminate the long-term need for dual-antiplatelet agents used for FDs and reduce associated risks, including thromboembolic events and bleeding, but also expedite incomplete and delayed healing in senior patients. As our understanding of vascular healing mechanisms improves, surface-modified implantable devices or adjunct treatments for aneurysm healing will mark the introduction of regenerative medicine into cerebrovascular disease.

### Study limitations

Although this represents a large prospective controlled study over a 5-year observation period with independent neurological adjudication and a core lab report on occlusion rates, the major limitations are the smaller sample size available in the senior population at various follow-up time points and small primary event rates, which may have impacted the results. A larger controlled cohort study may be helpful to confirm our observations.

## CONCLUSIONS AND RELEVANCE

Age > 65 and parent artery stenosis are related to an increased risk of major ipsilateral stroke in patients with intracranial aneurysms treated with a flow diverter. Age > 65 is also predictive of increased time to and incomplete healing. With the demographic shifts of patients treated, future technologies will have to focus on enhancing the cellular response required for expedited and improved healing.

## TRIAL PROTOCOL AND FUNDING

The SCENT Trial was sponsored and funded by Stryker Neurovascular and is registered at Clinicaltrials.gov (NCT01716117). Although the sponsor was involved in the design, collection, analysis, interpretation, and fact-checking of information, the authors independently made the content of this manuscript and its ultimate interpretation, along with the decision to submit it for publication.

## Data Availability

Yes, the clinical data referred to in the manuscript from the study will be available upon request.

